# Comparison of outcomes of second-line durvalumab plus tremelimumab and lenvatinib following first-line atezolizumab plus bevacizumab in unresectable hepatocellular carcinoma

**DOI:** 10.64898/2026.01.08.26343706

**Authors:** Chinatsu Nishioka, Yuki Tahata, Kazuki Maesaka, Machiko Kai, Kumiko Shirai, Kazuhiro Murai, Yuki Makino, Yoshinobu Saito, Yasutoshi Nozaki, Tasuku Nakabori, Hisashi Ishida, Takayuki Yakushijin, Sadaharu Iio, Nobuyuki Tatsumi, Kazuho Imanaka, Naruyasu Kakita, Ryotaro Sakamori, Atsushi Hosui, Masanori Miyazaki, Kengo Matsumoto, Masanori Nakahara, Yoshinori Doi, Mitsuru Sakakibara, Hayato Hikita, Takahiro Kodama, Tetsuo Takehara

## Abstract

**Background and Aim:** The optimal second-line therapy following first-line atezolizumab plus bevacizumab (Atezo/Beva) remains unascertained. This study compared second-line durvalumab plus tremelimumab (Dur/Tre) with lenvatinib (Len) after first-line Atezo/Beva in unresectable hepatocellular carcinoma (uHCC).

**Methods:** This prospectively registered cohort study analyzed patients with uHCC who received Dur/Tre (n = 14) or Len (n = 67) as second-line therapy after first-line Atezo/Beva. Tumor response was assessed by RECIST version 1.1. Progression-free survival (PFS), overall survival (OS), adverse events (AEs), and changes in the albumin–bilirubin (ALBI) score were compared.

**Results:** The objective response rate and disease control rate were 7.7% and 15.4% in the Dur/Tre group, and 23.3% and 76.7% in the Len group, respectively; median PFS was 1.7 vs. 4.2 months (*p* < 0.001) and median OS was 5.3 vs. 14.0 months (*p* = 0.047) in the Dur/Tre and Len groups, both significantly favoring Len. Multivariable analysis showed that Len treatment and neutrophil-to-lymphocyte ratio ≥3 were independent predictors of longer PFS and worse OS, respectively. Grade ≥3 AEs were more frequent with Len than with Dur/Tre (70.1% vs. 21.4%; *p* = 0.002). At week 4, the ALBI score did not worsen with Dur/Tre (−2.30 to −2.21; *p* = 0.318) but worsened with Len (−2.36 to −1.97; *p* < 0.001).

**Conclusions:** After first-line Atezo/Beva, second-line Len achieved superior disease control and longer survival than Dur/Tre. However, grade ≥ 3 AEs were more frequent with Len than with Dur/Tre, and careful management of AEs is essential when choosing therapy for individual patients.

## Introduction

Systemic therapy for unresectable hepatocellular carcinoma (HCC) commenced with sorafenib, which was approved as a first-line standard of care in 2009 ^[1,2]^. Subsequently, based on the REFLECT trial, lenvatinib (Len) was approved in 2018^[3]^. In 2020, the IMbrave150 trial established atezolizumab plus bevacizumab (Atezo/Beva) as the initial first-line regimen to incorporate an immune checkpoint inhibitor (ICI) ^[4]^. In 2022, following the HIMALAYA trial, durvalumab plus tremelimumab (Dur/Tre)—the first dual-immunotherapy regimen—was approved, and further expanded therapeutic options ^[5]^. Contemporary guidelines recommend Atezo/Beva or Dur/Tre as first-line therapy for patients who are eligible for ICI therapy ^[6–8]^. In 2025, nivolumab plus ipilimumab demonstrated a significant overall survival (OS) benefit, compared with Len or sorafenib, and further improvements in outcomes for patients with unresectable HCC are anticipated ^[9]^.

Atezo/Beva is an established first-line regimen for unresectable HCC ^[10]^. Treatment outcomes with Atezo/Beva included an objective response rate (ORR) of 22.0–28.2% and a disease control rate (DCR) of 69.6–70.6% ^[11,12]^. Although most patients experience disease progression during the Atezo/Beva treatment and transition to subsequent therapy, the optimal sequence of systemic treatments thereafter remains unclear ^[13]^. In this study, we evaluated the efficacy and safety of second-line Dur/Tre after first-line Atezo/Beva and compared these results with those of second-line Len after first-line Atezo/Beva.

## Methods

### Study Design

This multicenter, registry-based study was performed at The University of Osaka Hospital and 18 affiliated institutions of the Osaka Liver Forum. We aimed to analyze outcomes in patients who received Dur/Tre as second-line therapy after first-line Atezo/Beva (the Dur/Tre group) and to compare these outcomes with those of patients who received Len as second-line therapy after first-line Atezo/Beva (the Len group). Patient information was obtained from a prospectively registered observational cohort established by the participating centers and was subsequently evaluated for this investigation.

The study protocol was approved by The University of Osaka Hospital Ethical Review Board and the ethics committees of all participating institutions (UMIN000034611). Written informed consent was obtained from all participants at each participating institution. The data were accessed for research purposes between November 16, 2024, and September 3, 2025. After data collection, the authors did not have access to information that could identify individual participants.

### Study Population and Enrollment Period

In the Dur/Tre group, first-line Atezo/Beva treatment was initiated between October 2020 and January 2024; second-line Dur/Tre was started between April 2023 and June 2024 and was selected at the discretion of the attending physicians for each patient. In the Len group, 67 patients who received Len following first-line Atezo/Beva between October 2020 and January 2023, as reported previously ^[14]^, served as a historical control. By using this historical cohort, treatment-selection bias between Len and Dur/Tre after Atezo/Beva was considered to be minimized, because all patients in the Len group had been treated before Dur/Tre treatment became available. Finally, patients with unresectable HCC who received Dur/Tre or Len as second-line treatment following first-line Atezo/Beva were selected.

For both groups, the exclusion criteria were as follows: (1) initiation of Atezo/Beva as second-line or subsequent therapy; (2) inability to receive contrast-enhanced imaging for tumor response evaluation; (3) initiation of second-line Dur/Tre or Len >3 months after discontinuation of Atezo/Beva; (4) participation in a clinical trial; (5) Eastern Cooperative Oncology Group performance status of ≥ 2; and (6) observation period of < 4 weeks.

### Lenvatinib Administration

Len was administered orally once per day. According to the approved dosage in Japan ^[3,15,16]^, the initial dose was determined by body weight: 12 and 8 mg/day for patients ≥60 and <60 kg, respectively. Dose reductions or interruptions were allowed based on the severity of adverse events (AEs), in accordance with the prescribing recommendations.

### Durvalumab plus Tremelimumab Administration

Dur/Tre was administered according to the clinical trial protocol ^[5]^ as follows: 300 mg tremelimumab and 1500 mg durvalumab were administered on day 1, followed by 1500 mg durvalumab every 4 weeks. Dose interruptions were permitted based on the severity of AEs, in accordance with the prescribing recommendations.

### Outcome Measures

Progression-free survival (PFS) and OS were designated as the primary outcomes. The secondary outcomes were ORR, DCR, the incidence of treatment-related AEs, changes in the albumin–bilirubin (ALBI) score, and subsequent therapy.

### Treatment Evaluation

Based on the findings on contrast-enhanced computed tomography or magnetic resonance imaging, the tumor response was assessed using the Response Evaluation Criteria in Solid Tumors (RECIST) version 1.1 ^[17]^. Imaging evaluations were performed at baseline, at weeks 4 and 8 after treatment initiation, and every 8 weeks thereafter. AEs were evaluated and graded according to the Common Terminology Criteria for Adverse Events (CTCAE), version 5.0.

### Statistical Analysis

Clinical parameters were presented as counts and percentages for categorical variables, and as medians and ranges for continuous variables. Categorical variables were compared using the chi-square test or Fisher’s exact test, and continuous variables were compared with the Mann–Whitney *U* test. The PFS and OS were estimated using the Kaplan–Meier method, and differences between groups were assessed using the log-rank test. Independent prognostic factors in all patients associated with PFS and OS were identified using the Cox proportional hazards model. Based on prior literature ^[18–22]^, we prespecified the following covariates for inclusion in the multivariable analysis: treatment regimen, age, sex, etiology, Barcelona Clinic Liver Cancer stage, modified ALBI grades, serum α-fetoprotein level, des-γ-carboxy prothrombin (DCP) level, neutrophil-to-lymphocyte ratio (NLR), duration of prior Atezo/Beva and best response to prior Atezo/Beva treatment. Changes in the ALBI score within each group were compared using the Wilcoxon signed-rank test. A *p*-value < 0.05 was considered statistically significant. All statistical analyses were conducted using SPSS statistical software (version 29.0) for Windows (IBM Corp., Armonk, NY, USA).

## Results

### Patient Characteristics of Second-line Dur/Tre After First-line Atezo/Beva

The Dur/Tre group was developed by screening 35 patients who received Dur/Tre immediately after Atezo/Beva; after applying the exclusion criteria, 14 patients who had received first-line Atezo/Beva followed by second-line Dur/Tre were included in the analysis (**S1 Fig**). The median age was 73 years in this cohort including 9 men (64.3%). Child–Pugh class A and B were observed in 12 (85.7%) and 2 (14.3%) patients, respectively. Five or more intrahepatic tumors were present in 9 patients (64.3%), macrovascular invasion in 5 (35.7%), and extrahepatic metastasis in 6 (42.9%). Eight patients (57.1%) received prior Atezo/Beva for ≥3 months, and 9 (64.3%) achieved complete response (CR), partial response (PR), or stable disease (SD) on prior Atezo/Beva. All patients discontinued prior Atezo/Beva owing to disease progression; no discontinuations were attributed to AEs (**Table 1**). The median observation period was 5.6 (1.9–15.0) months.

**Table 1.** Patient Characteristics.

| Characteristic |  | Dur/Tre group<br>(n = 14) | Len group<br>(n = 67) | p value |
| --- | --- | --- | --- | --- |
| Age, years | Median (range) | 73 (31-88) | 75 (48-90) | 0.797 |
| Sex, n (%) | Male | 9 (64.3) | 56 (83.6) | 0.137 |
|  | Female | 5 (35.7) | 11 (16.4) |  |
| ECOG PS, n (%) | 0 | 12 (85.7) | 63 (94.0) | 0.276 |
|  | 1 | 2 (14.3) | 4 (6.0) |  |
| Etiology, n (%) | Viral | 4 (28.6) | 32 (47.8) | 0.244 |
|  | Non-viral | 10 (71.4) | 35 (52.2) |  |
| Maximum intrahepatic tumor size, mm | Median (range) | 37 (12-120) | 33 (0-168) | 0.546 |
| Intrahepatic tumor number, n (%) | ≤ 4 | 5 (35.7) | 21 (31.3) | 0.760 |
|  | ≥ 5 | 9 (64.3) | 46 (68.7) |  |
| Macrovascular invasion, n (%) | Absent | 9 (64.3) | 46 (68.7) | 0.760 |
|  | Present | 5 (35.7) | 21 (31.3) |  |
| Extrahepatic metastasis, n (%) | Absent | 8 (57.1) | 35 (52.2) | 0.777 |
|  | Present | 6 (42.9) | 32 (47.8) |  |
| BCLC stage, n (%) | A or B | 5 (35.7) | 24 (35.8) | 1.000 |
|  | C | 9 (64.3) | 43 (64.2) |  |
| Child-Pugh class, n (%) | A | 12 (85.7) | 58 (86.6) | 1.000 |
|  | B | 2 (14.3) | 9 (13.4) |  |
| mALBI grade, n (%) | 1 or 2a | 7 (50.0) | 38 (56.7) | 0.770 |
|  | 2b or 3 | 7 (50.0) | 29 (43.3) |  |
| AFP, ng/ml, n (%) | < 400 | 9 (64.3) | 48 (71.6) | 0.748 |
|  | ≥ 400 | 5 (35.7) | 19 (28.4) |  |
| DCP, mAU/ml, n (%) | < 400 | 6 (42.9) | 17 (26.2) | 0.330 |
|  | ≥ 400 | 8 (57.1) | 48 (73.8) |  |
| NLR, n (%) | < 3 | 4 (28.6) | 32 (49.2) | 0.238 |
|  | ≥ 3 | 10 (71.4) | 33 (50.8) |  |
| Duration of prior Atezo/Beva, months | < 3 | 6 (42.9) | 26 (38.8) | 0.773 |
|  | ≥ 3 | 8 (57.1) | 41 (61.2) |  |
| Best response to prior Atezo/Beva | CR/PR/SD | 9 (64.3) | 36 (53.7) | 0.562 |
|  | PD | 5 (35.7) | 31 (46.3) |  |
| Discontinuation reason of prior Atezo/Beva | PD | 14 (100.0) | 60 (89.6) | 0.345 |
|  | AE | 0 (0.0) | 7 (10.4) |  |

Abbreviations: Dur/Tre, durvalumab plus tremelimumab; Len, lenvatinib; ECOG PS, Eastern Cooperative Oncology Group performance status; BCLC, Barcelona Clinic Liver Cancer; mALBI, modified albumin-bilirubin; AFP, α-fetoprotein; DCP, des-gamma-carboxy prothrombin; NLR, neutrophil-to-lymphocyte ratio; Atezo/Beva, atezolizumab plus bevacizumab; CR, complete response; PR, partial response; SD, stable disease; PD, progressive disease; AE, adverse event

### Efficacy and Safety of Second-line Dur/Tre After First-line Atezo/Beva

According to the best therapeutic response assessed using RECIST version 1.1, the Dur/Tre group showed CR in 0 patients (0%), PR in 1 (7.7%), SD in 1 (7.7%), progressive disease (PD) in 11 (84.6%), and not evaluable (NE) in 1; this yielded an ORR of 7.7% and a DCR of 15.4% (**Table 2**). The median PFS was 1.7 (95% CI, 1.6–1.8) months and the median OS was 5.3 (95% CI, 0.3–10.4) months (**Fig 1A and 1B**).

**Fig 1.**
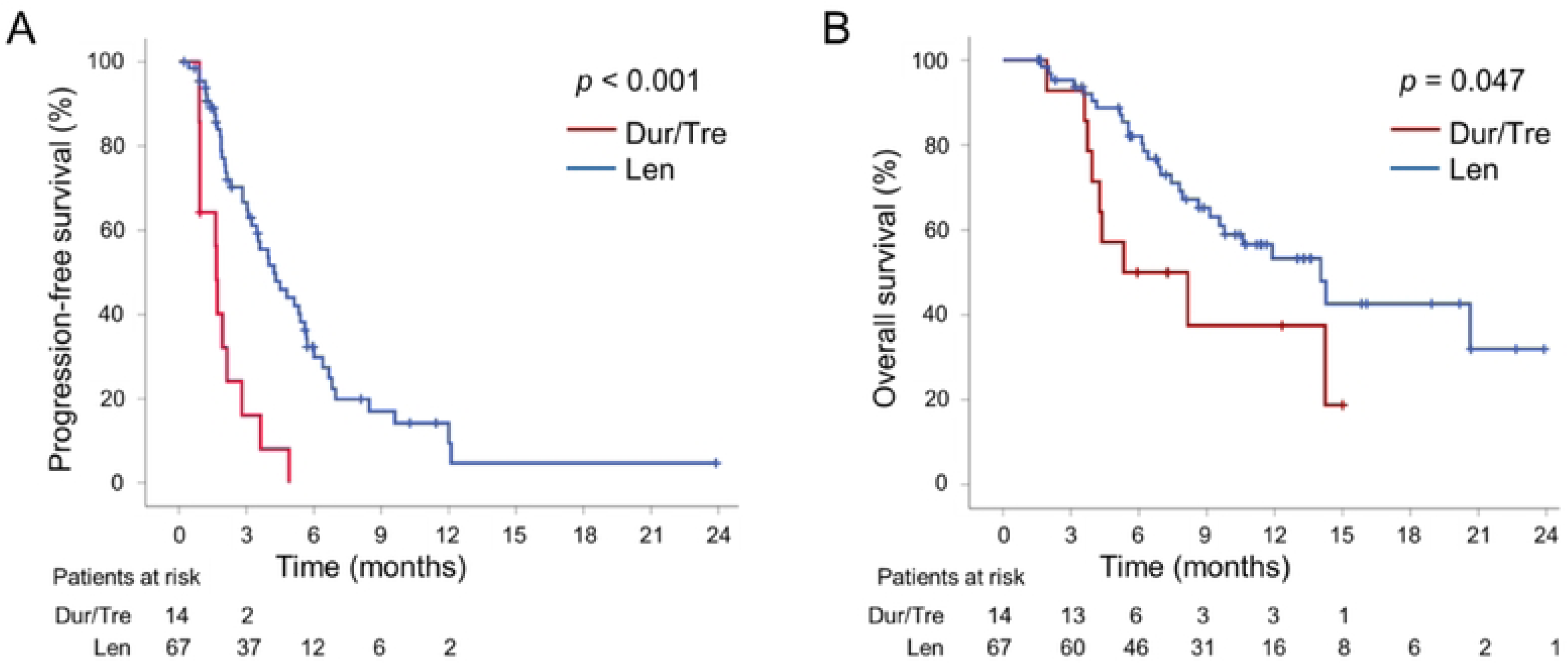
Comparisons of PFS (A) and OS (B) between the Dur/Tre and Len groups. Abbreviations: PFS, progression-free survival; OS, overall survival; Dur/Tre, durvalumab plus tremelimumab; Len, lenvatinib

**Table 2.**
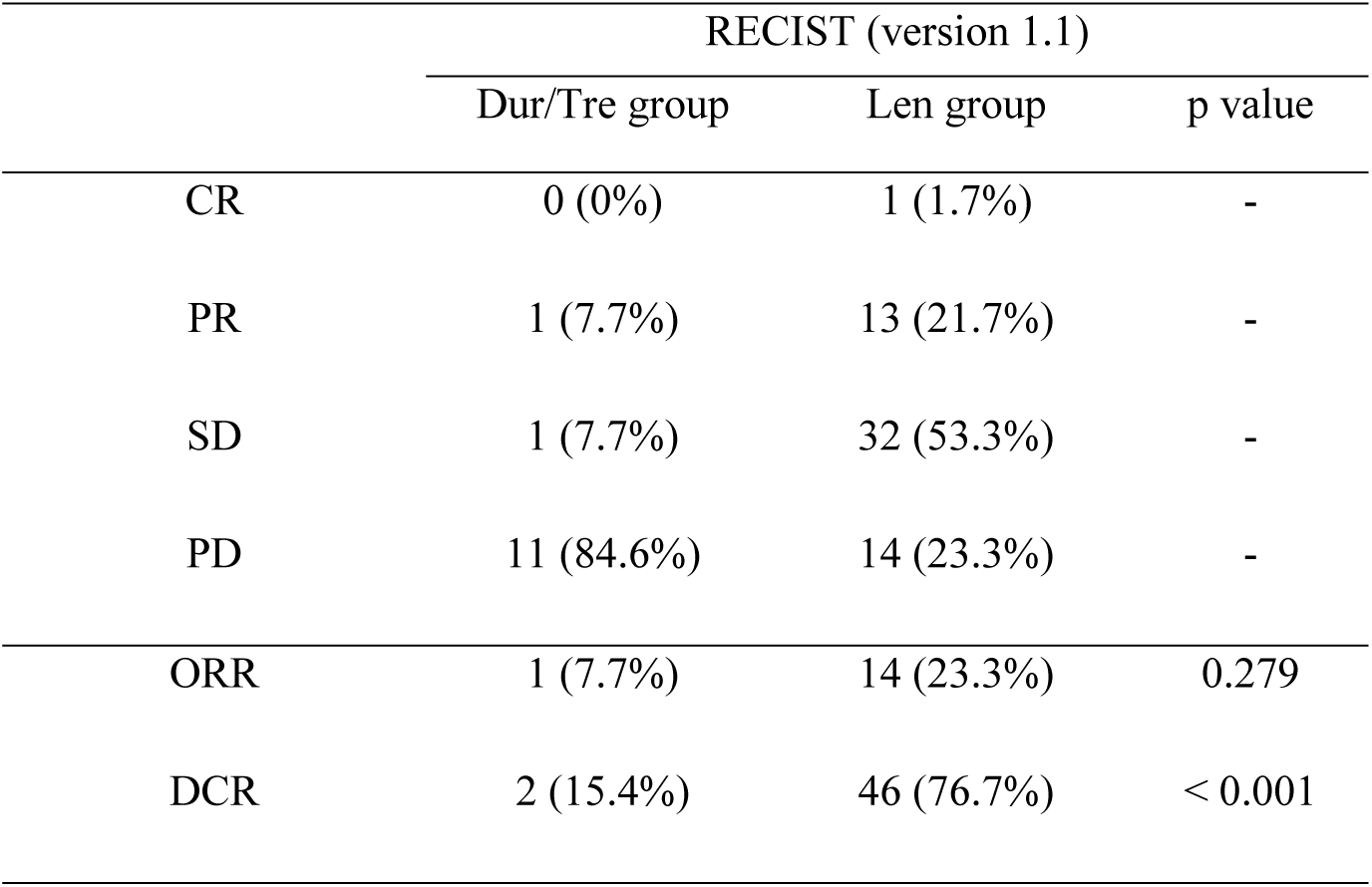
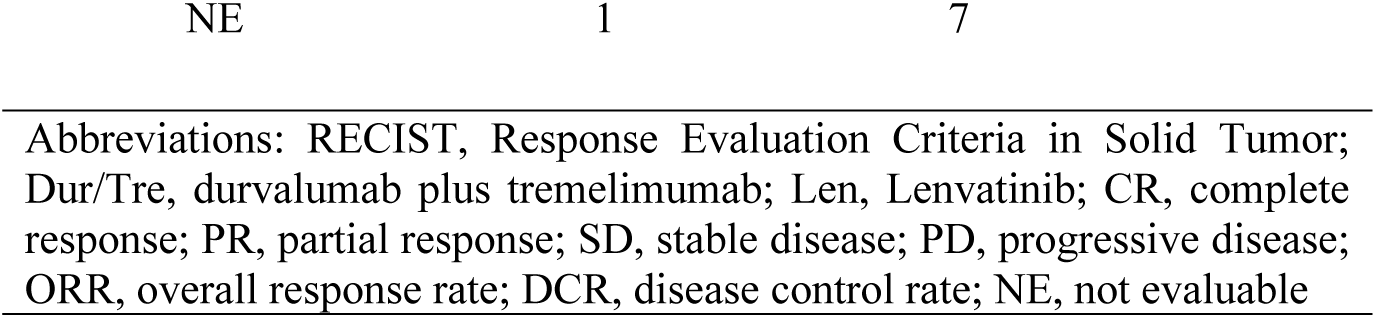
Best therapeutic response.

|  | RECIST (version 1.1) |  |  |
| --- | --- | --- | --- |
|  | Dur/Tre group | Len group | p value |
| CR | 0 (0%) | 1 (1.7%) | - |
| PR | 1 (7.7%) | 13 (21.7%) | - |
| SD | 1 (7.7%) | 32 (53.3%) | - |
| PD | 11 (84.6%) | 14 (23.3%) | - |
| ORR | 1 (7.7%) | 14 (23.3%) | 0.279 |
| DCR | 2 (15.4%) | 46 (76.7%) | < 0.001 |

Abbreviations: RECIST, Response Evaluation Criteria in Solid Tumor; Dur/Tre, durvalumab plus tremelimumab; Len, Lenvatinib; CR, complete response; PR, partial response; SD, stable disease; PD, progressive disease; ORR, overall response rate; DCR, disease control rate; NE, not evaluable

The incidence of any-grade and grade ≥3 AEs was 78.6% and 21.4%, respectively (**Table 3**). The most frequent any-grade AEs were rash (28.6%), increased aspartate aminotransferase or alanine aminotransferase (21.4%) level, and hypothyroidism (14.3%). Grade ≥3 AEs occurred in 3 patients (21.4%): rash in 1 (7.1%), hepatic failure in 1 (7.1%), and tumor bleeding in 1 (7.1%). Systemic corticosteroids were administered for immune-related adverse events (irAEs) in 2 patients (14.3%)—rash in 1 (7.1%) and colitis in 1 (7.1%).

**Table 3.** Adverse Events.

|  | Any-grade; n (%) |  |  | ≥ Grade 3; n (%) |  |  |
| --- | --- | --- | --- | --- | --- | --- |
|  | Dur/Tre group | Len group | p value | Dur/Tre group | Len group | p value |
| Any adverse event | 11 (78.6) | 66 (98.5) | 0.015 | 3 (21.4) | 47 (70.1) | 0.002 |
| Rash | 4 (28.6) | 10 (14.9) | 0.249 | 1 (7.1) | 2 (3.0) | 0.439 |
| Increased AST or ALT | 3 (21.4) | 12 (17.9) | 0.716 | 0 (0.0) | 3 (4.5) | 1.000 |
| Hypothyroidism | 2 (14.3) | 25 (37.3) | 0.125 | 0 (0.0) | 0 (0.0) | NA |
| Colitis / diarrhea | 1 (7.1) | 10 (14.9) | 0.679 | 0 (0.0) | 1 (1.5) | 1.000 |
| Adrenal insufficiency | 1 (7.1) | 0 (0.0) | 0.173 | 0 (0.0) | 0 (0.0) | NA |
| Fatigue | 1 (7.1) | 38 (56.7) | < 0.001 | 0 (0.0) | 8 (11.9) | 0.339 |
| Proteinuria | 1 (7.1) | 37 (55.2) | < 0.001 | 0 (0.0) | 20 (29.9) | 0.017 |
| Decreased appetite | 1 (7.1) | 35 (52.2) | 0.002 | 0 (0.0) | 6 (9.0) | 0.583 |
| Hypertension | 0 (0.0) | 32 (47.8) | < 0.001 | 0 (0.0) | 9 (13.4) | 0.347 |
| Thrombocytopenia | 0 (0.0) | 17 (25.4) | 0.034 | 0 (0.0) | 3 (4.5) | 1.000 |
| Hand-foot-skin reaction | 0 (0.0) | 20 (29.9) | 0.017 | 0 (0.0) | 2 (3.0) | 1.000 |
| Liver failure | 1 (7.1) | 0 (0.0) | 0.173 | 1 (7.1) | 0 (0.0) | 0.173 |
| Bleeding | 1 (7.1) | 3 (4.5) | 0.539 | 1 (7.1) | 1 (1.5) | 0.318 |

Abbreviations: Dur/Tre, durvalumab plus tremelimumab; Len, Lenvatinib; AST, aspartate aminotransferase; ALT, alanine aminotransferase; NA, not applicable

### Comparative Efficacy and Safety of Second-line Dur/Tre and Len After First-line Atezo/Beva

In the Len group, patient characteristics, efficacy, and safety were previously reported ^[14]^. There were no significant between-group differences in baseline characteristics (**Table 1**) or the ORR (the Dur/Tre group, 7.7% vs. the Len group, 23.3%; *p* = 0.279), whereas the DCR was significantly higher in the Len group than in the Dur/Tre group (15.4% vs. 76.7%; *p* < 0.001; **Table 2**). The median PFS was 1.7 (95% CI, 1.6–1.8) and 4.2 (95% CI, 2.9–5.6) months in the Dur/Tre and Len groups, respectively, and favored Len (*p* < 0.001). The median OS was 5.3 (95% CI, 0.3–10.4) and 14.0 (95% CI, 9.8–18.2) months in the Dur/Tre and Len groups, respectively, favoring Len (*p* = 0.047; **Fig 1A and 1B**).

The incidences of any-grade and grade ≥3 AEs were significantly higher in the Len group than in the Dur/Tre group (**Table 3**). Among any-grade AEs, fatigue, proteinuria, decreased appetite, hypertension, thrombocytopenia, and hand-foot skin reaction were significantly more common in the Len group. Proteinuria was also significantly more frequent in the Len group among grade ≥3 AEs. Treatment discontinuation owing to AEs occurred in 2 patients (14.3%) in the Dur/Tre group and 18 patients (26.9%) in the Len group, without significant between-group differences (*p* = 0.499).

### Clinical Factors Associated With PFS and OS in the Overall Cohort

The pretreatment clinical variables were analyzed to identify risk factors associated with shorter PFS and OS in the overall cohort. In multivariable analysis, including treatment regimen, only Len treatment (HR 0.271, 95% CI 0.129–0.567; *p* < 0.001) was identified as an independent factor that was associated with prolonged PFS (**Table 4**). In multivariable analysis using the same covariates as for PFS, only elevated NLR (≥3; HR 2.394, 95% CI 1.067–5.375; *p* = 0.034) was independently associated with OS (**Table 5**). Median OS was shorter in patients with NLR ≥3 than in those with NLR <3 (8.6 vs. 20.6 months; **S2 Fig**).

**Table 4.** Multivariable analyses of PFS-related factors.

| Viabiles | Category | Multivariable analysis | p value |
| --- | --- | --- | --- |
|  |  | Hazard ratio<sup>†</sup><br>(95% CI) |  |
| Treatment | Dur/Tre | 1 | < 0.001 |
|  | Len | 0.271 (0.129-0.567) |  |
| Age, years | < 75 | 1 | 0.224 |
| | $\geq 75$ | 1.469 (0.790-2.733) | |
| Sex | Male | 1 | 0.399 |
|  | Female | 1.356 (0.668-2.750) |  |
| Etiology | Viral | 1 | 0.492 |
|  | Non-viral | 1.249 (0.663-2.353) |  |
| BCLC stage | A or B | 1 | 0.213 |
|  | C | 1.635 (0.755-3.543) |  |
| mALBI grade | 1 or 2a | 1 | 0.680 |
|  | 2b or 3 | 1.133 (0.626-2.050) |  |
| AFP, ng/mL | < 400 | 1 | 0.320 |
|  | ≥ 400 | 1.426 (0.709-2.867) |  |
| DCP, mAU/ml | < 400 | 1 | 0.983 |
|  | ≥ 400 | 1.007 (0.527-1.923) |  |
| NLR | < 3 | 1 | 0.134 |
|  | ≥ 3 | 1.603 (0.865-2.971) |  |
| Duration of prior Atezo/Beva, months | < 3 | 1 | 0.608 |
|  | ≥ 3 | 0.825 (0.395-1.722) |  |
| Best response to prior Atezo/Beva | CR/PR/SD | 1 | 0.692 |
|  | PD | 0.851 (0.382-1.896) |  |

Abbreviations: CI, confidence interval; PFS, progression-free survival; Dur/Tre, durvalumab plus tremelimumab; Len, lenvatinib; BCLC, Barcelona Clinic Liver Cancer; mALBI, modified albumin-bilirubin; AFP, α-fetoprotein; DCP, des-gamma-carboxy prothrombin; NLR, neutrophil-to-lymphocyte ratio; Atezo/Beva, atezolizumab plus bevacizumab
†Adjusted for treatment, age, sex, etiology, BCLC stage, mALBI grade, AFP, DCP, NLR, duration of prior Atezo/Beva, best response to prior Atezo/Beva

**Table 5.** Multivariable analyses of OS-related factors.

| Viabiles | Category | Multivariable analysis | p value |
| --- | --- | --- | --- |
|  |  | Hazard ratio<sup>†</sup><br>(95% CI) |  |
| Treatment | Dur/Tre | 1 | 0.103 |
|  | Len | 0.431 (0.157-1.184) |  |
| Age, years | < 75 | 1 | 0.551 |
|  | ≥ 75 | 1.311 (0.538-3.196) |  |
| Sex | Male | 1 | 0.192 |
|  | Female | 1.897 (0.726-4.960) |  |
| Etiology | Viral | 1 | 0.971 |
|  | Non-viral | 1.016 (0.431-2.397) |  |
| BCLC stage | A or B | 1 | 0.549 |
|  | C | 1.458 (0.425-5.005) |  |
| mALBI grade | 1 or 2a | 1 | 0.242 |
|  | 2b or 3 | 1.557 (0.741-3.272) |  |
| AFP, ng/mL | < 400 | 1 | 0.698 |
|  | ≥ 400 | 1.165 (0.538-2.525) |  |
| DCP, mAU/ml | < 400 | 1 | 0.181 |
|  | ≥ 400 | 1.943 (0.734-5.139) |  |
| NLR | < 3 | 1 | 0.034 |
|  | ≥ 3 | 2.394 (1.067-5.375) |  |
| Duration of prior Atezo/Beva, months | < 3 | 1 | 0.947 |
|  | ≥ 3 | 0.966 (0.354-2.637) |  |
| Best response to prior Atezo/Beva | CR/PR/SD | 1 | 0.135 |
|  | PD | 2.480 (0.753-8.168) |  |

Abbreviations: CI, confidence interval; OS, overall survival; Dur/Tre, durvalumab plus tremelimumab; Len, lenvatinib; BCLC, Barcelona Clinic Liver Cancer; mALBI, modified albumin-bilirubin; AFP, α-fetoprotein; DCP, des-gamma-carboxy prothrombin; NLR, neutrophil-to-lymphocyte ratio; Atezo/Beva, atezolizumab plus bevacizumab
<sup>†</sup>Adjusted for treatment, age, sex, etiology, BCLC stage, mALBI grade, AFP, DCP, NLR, duration of prior Atezo/Beva, best response to prior Atezo/Beva

### Changes in the ALBI Score

The median ALBI score in the Dur/Tre group was −2.30 (IQR, −2.51 to −2.00) at treatment initiation and −2.21 (IQR, −2.50 to −1.92) at week 4, without significant deterioration (*p* = 0.318). In the Len group, the median ALBI score was −2.36 (IQR, −2.62 to −2.07) at initiation and worsened to −1.97 (IQR, −2.27 to −1.61) at week 4 (*p* < 0.001; **Fig 2**).

**Fig 2.**
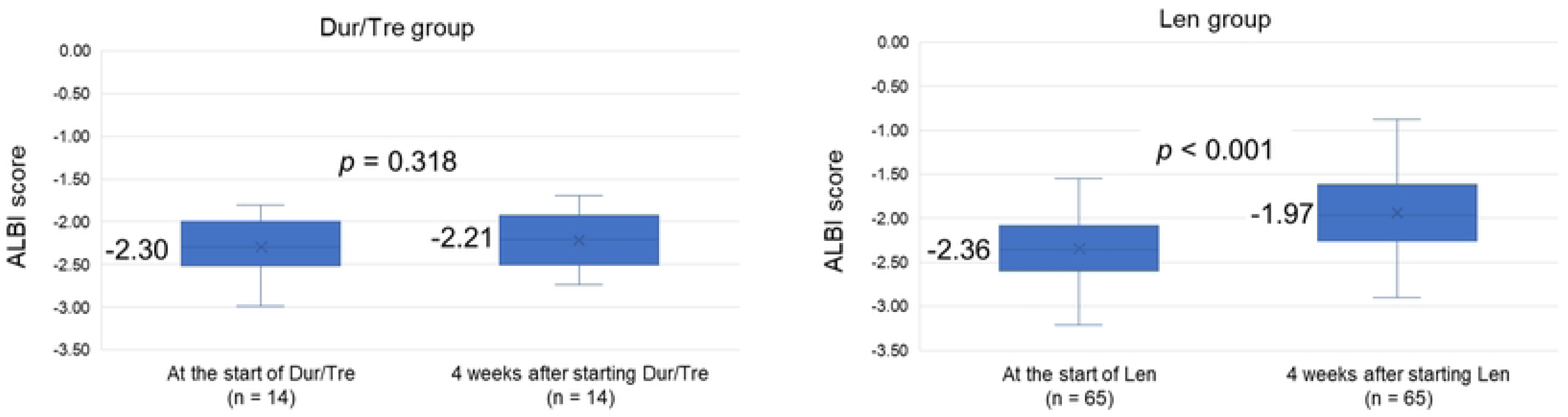
Within-group changes in the ALBI score from treatment initiation to week 4 in the Dur/Tre and Len groups. Abbreviations: ALBI, albumin–bilirubin; Dur/Tre, durvalumab plus tremelimumab; Len, lenvatinib

### Subsequent Therapy

In the Dur/Tre group, after the discontinuation of Dur/Tre, 10 of the 14 patients (71.4%) received subsequent therapy whereas the remaining 4 (28.6%) received best supportive care. In the Len group, after discontinuation of Len, 48 of the 66 patients (72.7%) received subsequent therapy and the remaining 18 (27.3%) received best supportive care; 1 patient remained on Len at the data-cutoff timepoint (**S1 Table**). There was no significant between-group difference in the proportion that received subsequent therapy (*p* = 1.000).

## Discussion

This multicenter collaborative study of patients with unresectable HCC to compare the efficacy and safety of second-line Dur/Tre versus Len following first-line Atezo/Beva showed that the Len group had significantly better PFS, OS (Fig 1A and 1B), and DCR (Table 2) than the Dur/Tre group. However, severe AEs were significantly more frequent in the Len group than in the Dur/Tre group (Table 3). Unlike the Dur/Tre group, the Len group exhibited a significant worsening of the ALBI score after treatment initiation (Fig 2). To the best of our knowledge, this is the first report of a direct comparison of second-line Dur/Tre and Len after first-line Atezo/Beva.

On second-line therapy after first-line Atezo/Beva, the Len group showed superior PFS, OS, and DCR compared with the Dur/Tre group. Maesaka et al. recently compared first-line Len with second-line Len after Atezo/Beva and reported comparable efficacy in the two groups ^[14]^. Other real-world studies have reported stable responses and favorable survival with second-line Len after Atezo/Beva ^[23–25]^; therefore, Len can be expected to be effective as second-line therapy after Atezo/Beva. Pharmacologically, prolonged programmed death-ligand 1 (PD-L1) blockade on CD8+ T cells induced by Atezo/Beva may potentiate the antitumor activity of subsequent Len treatment ^[26,27]^. In contrast, several reports suggest reduced effectiveness of Dur/Tre after Atezo/Beva ^[19,28]^. Fujiwara et al. identified an independent association of prior Atezo/Beva with shorter OS (HR 27.38, 95% CI 1.92–391.50; *p* = 0.01) and shorter PFS (HR 6.25, 95% CI 1.16–33.71; *p* = 0.03) in multivariable Cox analyses of Dur/Tre-treated patients ^[19]^. Mori et al. reported that, compared with first-line Dur/Tre and later-line Dur/Tre without prior Atezo/Beva, later-line Dur/Tre with prior Atezo/Beva showed significantly worse ORR/DCR and PFS ^[28]^. The potential mechanisms underlying the attenuated Dur/Tre efficacy after Atezo/Beva include resistance to programmed death 1 (PD-1)/PD-L1 blockade with T-cell exhaustion ^[29]^, diminished T-cell priming owing to anti-VEGF resistance ^[30]^, and rebound VEGF-mediated immunosuppression after bevacizumab ^[31]^. Nevertheless, research in large prospective datasets under uniform conditions regarding Dur/Tre after Atezo/Beva remain scarce, and this necessitates further investigation.

Regarding hepatic reserve, the Dur/Tre group maintained the ALBI score from treatment initiation to week 4, whereas the Len group showed worsening ALBI (Fig 2). Miura et al. demonstrated preservation of liver function before and after Dur/Tre in patients with HCC who were treated with Dur/Tre following Atezo/Beva ^[32]^. Tamaki et al. reported that Dur/Tre can maintain liver function throughout treatment even in later-line settings and in patients with impaired baseline function ^[33]^. Accordingly, Dur/Tre is considered likely to preserve liver function. In our cohort, such preservation likely contributed to a transition rate to subsequent therapy that was comparable to that in the Len group (S1 Table). In contrast, Len caused early decline in liver function in a subset of patients ^[34–36]^, which is consistent with our observations. From the standpoint of preserving liver function, Dur/Tre may be considered a reasonable option even after Atezo/Beva.

Regarding safety, the incidence of AEs was significantly higher in the Len group, wherein 98.5% had at least one AE and 70.1% had grade ≥3 AEs; fatigue, proteinuria, decreased appetite, and hypertension were common (Table 3). Via VEGF/VEGFR inhibition, Len injures the vascular endothelium, glomerular endothelium/podocytes, and the thyroid capillary network, thereby leading to hypertension, proteinuria, and hypothyroidism; furthermore, fatigue and anorexia have a multifactorial pathogenesis ^[37–39]^. The AE profile observed in our Len cohort was broadly consistent with prior reports of patients treated with Len after Atezo/Beva ^[22,26,27]^. In the Dur/Tre group, although rash and hepatic dysfunction were relatively frequent, the overall incidence of AEs—both any-grade and grade ≥3—was lower as compared with the Len group. With Dur/Tre, the activation of tumor- and self-reactive T cells can systemically injure normal tissues and induce T-cell–mediated irAEs, such as dermatitis and autoimmune hepatitis–like inflammation ^[40,41]^. The AE profile in our Dur/Tre cohort showed a similar pattern to that reported by Miura et al. for Dur/Tre administered after Atezo/Beva. Overall, the frequency of severe AEs with Dur/Tre after Atezo/Beva was low, which supports an acceptable safety profile.

This study has some limitations. First, the sample size was limited, especially in the Dur/Tre group. Second, clinical differences in treatment availability and practice patterns between Dur/Tre and Len cannot be fully excluded. To mitigate this real-world bias, in this study, the Len group was constituted from a historical cohort that was treated before Dur/Tre became available; nevertheless, as the patients were not prospectively allocated to either regimen, residual selection bias cannot be ruled out.

In conclusion, as second-line therapy after Atezo/Beva, Len showed better treatment outcomes than Dur/Tre; however, severe AEs were more frequent with Len. Careful AE management is therefore essential when selecting second-line therapy in patients with unresectable HCC after Atezo/Beva.

## Data Availability

All relevant data are within the manuscript and its Supporting Information files.

## Acknowledgments

The authors are grateful to all the physicians who contributed to this study.

## Ethics approval statement

The study protocol was reviewed and approved by the Institutional Review Board or Ethics Committee at each participating center (UMIN 000034611). All procedures were conducted in accordance with the principles of the Declaration of Helsinki. Written informed consent was obtained from all patients who were enrolled in the registry cohort.

### Abbreviations

AE: adverse event;
ALBI: albumin–bilirubin;
Atezo/Beva: atezolizumab plus bevacizumab;
CI: confidence interval
CR: complete response;
CTCAE: Common Terminology Criteria for Adverse Events;
DCR: disease control rate;
Dur/Tre: durvalumab plus tremelimumab;
HCC: hepatocellular carcinoma;
ICI: immune checkpoint inhibitor;
IQR: interquartile range;
irAE: immune-related adverse event;
Len: lenvatinib;
ORR: objective response rate;
OS: overall survival;
PD: progressive disease;
PD-1: programmed death 1;
PD-L1: programmed death-ligand 1;
PFS: progression-free survival;
PR: partial response;
RECIST: Response Evaluation Criteria in Solid Tumors;
SD: stable disease

## Supporting information captions

**S1 Fig.** Flowchart of the participant enrollment

Abbreviations: Dur/Tre, durvalumab plus tremelimumab; Len, lenvatinib; Atezo/Beva, atezolizumab plus bevacizumab; PS, performance status

**S2 Fig.** Comparisons of OS between patients with NLR ≥3 and <3 Abbreviations: OS, overall survival; NLR, neutrophil-to-lymphocyte ratio

